# A Closed-Loop Digital Twin Framework for Automated Control of Extracorporeal Membrane Oxygenation: Toward Physiology-Aware Decision Support in Critical Care

**DOI:** 10.64898/2026.01.01.26343307

**Authors:** Amin Ramezani, Till Seefeldt, Steve P. Keller, Amy E. Hackmann, Asishana Osho, S. Alireza Rabi, Farhad R. Nezami

## Abstract

Extracorporeal membrane oxygenation (ECMO) is a life-saving therapy for severe cardiopulmonary failure, yet its management remains highly manual, experience-dependent, and vulnerable to delayed or suboptimal adjustments in rapidly evolving clinical states. Clinicians must continuously balance oxygen delivery, carbon dioxide removal, and hemodynamic stability using sparse, intermittently sampled physiologic data, creating substantial cognitive and operational burden in high-acuity intensive care settings. Here, we present a closed-loop digital twin framework for physiology-aware ECMO control that integrates a mechanistic cardiopulmonary model with constrained model predictive control to enable real-time, adaptive decision support. The digital twin explicitly represents patient-specific interactions between cardiac function, pulmonary gas exchange, and extracorporeal support, allowing continuous estimation of physiologic state and forward prediction under changing clinical conditions.

The framework was evaluated in silico across representative clinical scenarios, including acute respiratory distress syndrome and cardiogenic shock, capturing distinct pathophysiologic regimes encountered in ECMO practice. Simulation results demonstrate that the proposed approach maintains arterial oxygenation and carbon dioxide targets while preserving hemodynamic stability and respecting clinically meaningful safety constraints. Compared with static or heuristic control strategies, the digital twin–driven controller exhibits improved robustness to disturbances and parameter uncertainty, supporting consistent performance across heterogeneous disease states. This study establishes a foundation for digital twin–based ECMO decision-support systems that augment clinician oversight with transparent, physiology-grounded intelligence. By enabling predictive, interpretable, and adaptable control in silico, this work advances the broader vision of digital medicine in critical care and provides a scalable pathway toward safer, data-informed extracorporeal support.

“Digital Twin-based Model Predictive Control for Extracorporeal Membrane Oxygenation (ECMO).” A mechanistic 0D cardiopulmonary digital twin integrates patient physiology with oxygenator diffusion dynamics. This model enables an MPC controller to autonomously titrate ECMO settings (Q_b_, sweep gas, F_i_O_2_) to maintain precise physiological targets in simulated ARDS and Cardiogenic Shock cohorts.

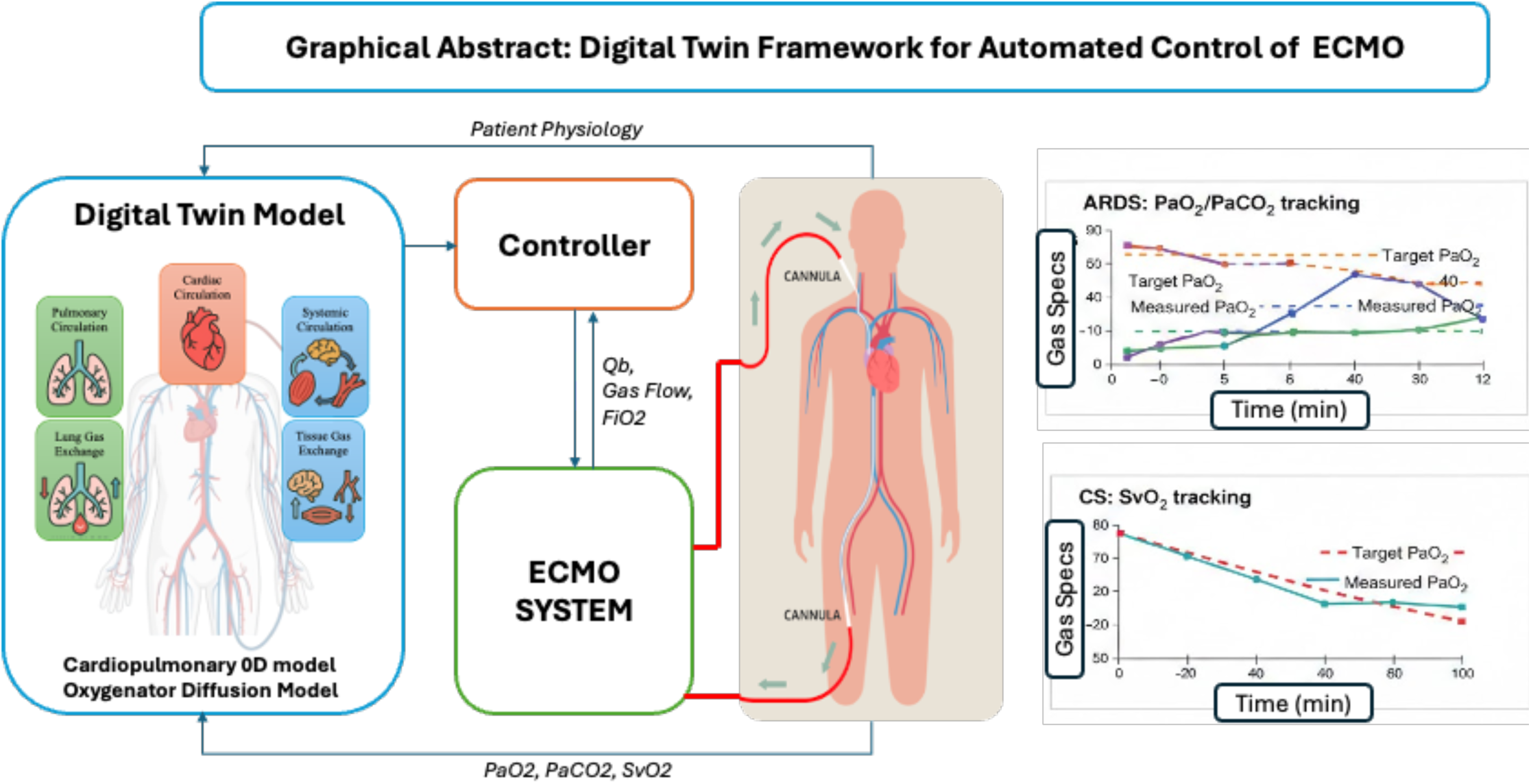

## 1. Introduction

Extracorporeal membrane oxygenation (ECMO) is a life-sustaining therapy for patients with refractory respiratory or circulatory failure, providing extracorporeal gas exchange and circulatory support via a membrane oxygenator before reinfusion into the systemic circulation [1–3]. Over the past decade, ECMO utilization has expanded substantially owing to advances in cannulation strategies, oxygenator efficiency, biocompatible materials, and standardized clinical protocols [4–6]. Consequently, ECMO has evolved from a rescue therapy of last resort into a central component of modern critical care, supporting a wide spectrum of respiratory and cardiac failure phenotypes, most prominently severe acute respiratory distress syndrome (ARDS) and cardiogenic shock (CS) [7, 8]. Despite these technological advances, ECMO management remains largely manual. Clinicians adjust extracorporeal blood flow (Qb), sweep gas flow (Qg), and sweep gas oxygen fraction (FiO₂) based on intermittent blood gas sampling and evolving metabolic demands [9–11]. This inherently reactive workflow applies corrective interventions only after deviations in arterial oxygen (PaO₂), carbon dioxide (PaCO₂), or mixed venous oxygen saturation (SvO₂) are detected. Such delays expose patients to prolonged hypoxemia, hyperoxemia, or hypercapnia, which are associated with neurological injury, multiorgan dysfunction, and increased mortality, particularly in ARDS populations [12, 13].

Digital medicine in critical care increasingly demands computational systems capable of continuously assimilating physiologic data, predicting near-term trajectories, and supporting safe, time-critical interventions. A clinical digital twin may be defined as a patient-linked mechanistic model that operates alongside bedside care to provide forward-looking, actionable predictions under candidate therapeutic adjustments. ECMO represents a particularly compelling use case for digital-twin deployment due to its tightly coupled, nonlinear gas-exchange and circulatory dynamics, combined with routine reliance on intermittent measurements and clinician experience. Importantly, ECMO control objectives are phenotype dependent: respiratory-dominant failure (e.g., ARDS on VV-ECMO) is governed by rapid gas-exchange dynamics and shunt nonlinearities, whereas hemodynamic-dominant failure (e.g., CS on VA-ECMO) is constrained by slower venous oxygen kinetics and circulatory transport delays. These distinctions motivate a pathology-aware digital twin and control strategy rather than a single universal control law. ARDS remains the most common indication for veno-venous ECMO, with rising global prevalence driven by infectious and noninfectious etiologies [14, 15]. Its hallmark features, reduced lung compliance, elevated shunt fraction, dynamic airway resistance, and pulmonary vascular dysfunction, produce highly nonlinear and time-varying gas-exchange behavior that challenges conventional control strategies [16–19]. In contrast, cardiogenic shock is characterized by impaired myocardial contractility, reduced cardiac output, elevated filling pressures, and systemic hypoperfusion, frequently necessitating veno-arterial ECMO for circulatory stabilization and end-organ support [20–22]. These contrasting respiratory-dominant and hemodynamic-dominant failure mechanisms under ECMO support are illustrated in Figure 1.

**Figure 1:**
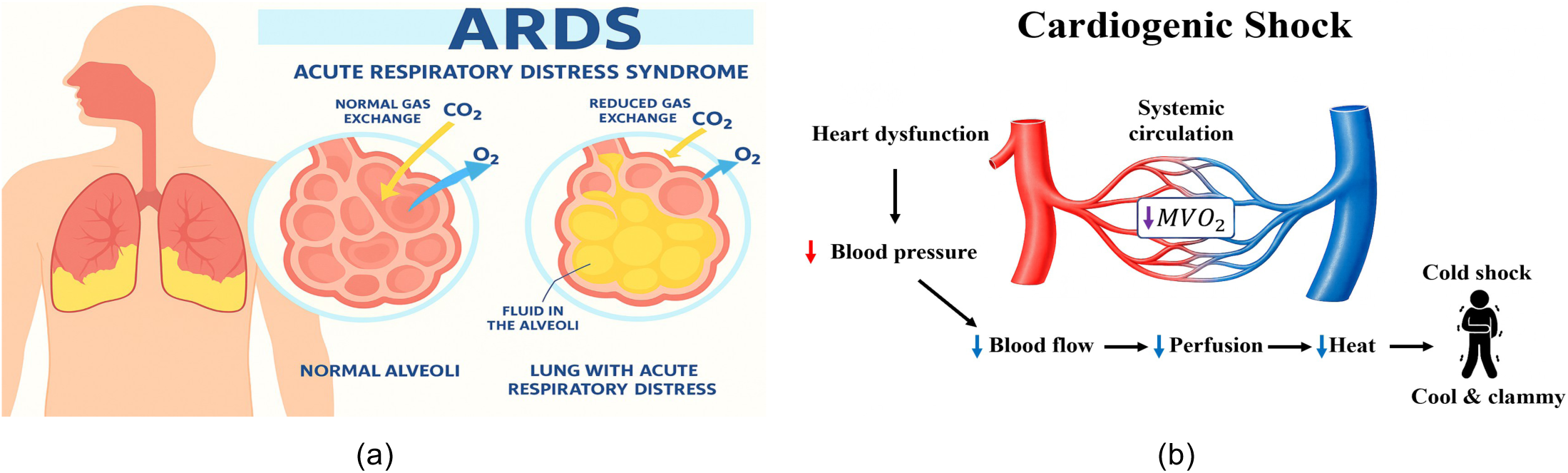
Contrasting respiratory-dominant (ARDS) versus hemodynamic-dominant (cardiogenic shock) failure mechanisms under ECMO support. ARDS: Respiratory-dominant failure with impaired alveolar gas exchange (LEFT). Cardiogenic shock: Hemodynamic-dominant failure with reduced systemic perfusion and oxygen delivery (RIGHT).

These indications present fundamentally different control challenges. ARDS demands rapid, precise regulation of PaO₂ and PaCO₂ in the presence of unstable pulmonary mechanics, whereas cardiogenic shock requires stabilization of SvO₂ and systemic perfusion under slower venous oxygen dynamics and constrained cardiac output. Yet in both scenarios, clinicians must manually tune the same extracorporeal actuators under extreme time pressure and physiologic uncertainty. This mismatch between system complexity and manual control capacity remains a central limitation of contemporary ECMO therapy. Consequently, automated closed-loop ECMO control has emerged as a promising paradigm for continuous, physiology-aware extracorporeal support [23–25]. Prior studies have explored proportional–integral, fuzzy logic, adaptive, and extremum-seeking control approaches for oxygen and carbon dioxide regulation [26–29].

While these efforts demonstrate feasibility, most rely on simplified gas-exchange representations and do not explicitly incorporate the full cardiopulmonary circulation, limiting generalizability across ECMO indications and hemodynamic instability. Digital twin modeling provides a principled framework to address these limitations by unifying physiological modeling, gas-exchange dynamics, and advanced control within a predictive architecture [30–32]. Lumped-parameter cardiopulmonary models effectively capture coupled systemic–pulmonary interactions at clinically relevant time scales [33–35], and membrane oxygenator behavior has been rigorously characterized using diffusion-based transport and nonlinear hemoglobin kinetics [36–38]. However, integration of these components into a fully closed-loop, pathology-aware ECMO control system remains limited.

Model predictive control (MPC) offers a natural control paradigm for such constrained, multivariable physiological systems [39–42] and has achieved clinical translation in domains such as anesthesia and metabolic regulation [43]. Nevertheless, its application to a fully integrated ECMO digital twin capable of simultaneous regulation of PaO₂, PaCO₂, and SvO₂ across distinct ECMO pathologies remains largely unexplored. Specifically, this work: Integrates a lumped-parameter cardiopulmonary circulation model with a nonlinear diffusion-based membrane oxygenator into a unified ECMO digital twin, Develops pathology-specific multivariable MPC strategies for ARDS and cardiogenic shock, Demonstrates simultaneous regulation of PaO₂, PaCO₂, and SvO₂ under realistic metabolic disturbances while enforcing actuator and safety constraints, Establishes an in-silico testbed for safety evaluation, controller tuning, and future patient-specific adaptation of autonomy-assisted extracorporeal support.

In this study, we develop a physiology-grounded, closed-loop digital twin framework for ECMO control using multivariable MPC. By coupling real-time physiological prediction with constrained control, the framework supports robust gas-exchange regulation while enabling in-silico safety testing, controller optimization, and future integration with bedside monitoring systems. From a digital medicine perspective, this work reframes ECMO as a continuously optimized, data-driven therapy rather than a manually adjusted support modality, advancing predictive, transparent, and interpretable decision support for high-acuity critical care.

## 2. System Overview

The proposed closed-loop digital twin framework unifies cardiopulmonary physiology, extracorporeal gas exchange, and constrained multivariable control within a single real-time predictive architecture. Rather than treating ECMO as an externally tuned support device, the framework embeds the extracorporeal circuit directly within a mechanistic representation of patient cardiovascular and respiratory dynamics, enabling continuous physiology-aware regulation of artificial gas exchange. The system comprises four tightly integrated components: (i) a lumped-parameter cardiopulmonary circulation model; (ii) a nonlinear diffusion-based membrane oxygenator model; (iii) a pathology modulation layer that reconfigures model parameters to represent disease-specific physiology; and (iv) multivariable model predictive controllers (MPCs) that compute optimal ECMO control actions in real time. Together, these elements form a closed feedback loop in which physiological state prediction and control optimization evolve synchronously.

The cardiopulmonary circulation model provides a coupled representation of ventricular function, pulmonary blood flow, systemic perfusion, and gas transport between lung, blood, and tissue compartments. This model serves as the predictive backbone of the digital twin, yielding state estimates of arterial oxygen tension (PaO₂), arterial carbon dioxide tension (PaCO₂), and mixed venous oxygen saturation (SvO₂). The membrane oxygenator is embedded directly within the circulation model, allowing extracorporeal gas exchange to be described using first-principles diffusion and hemoglobin dissociation kinetics rather than empirical transfer coefficients alone.

Pathology emulation is implemented as an intermediate layer that transforms baseline physiological parameters into disease-specific operating regimes. In acute respiratory distress syndrome, reduced lung compliance, elevated airway resistance, and increased intrapulmonary shunting reproduce the nonlinear gas-exchange instability characteristic of ARDS. In cardiogenic shock, depressed ventricular contractility, elevated systemic afterload, and constrained cardiac output produce slow venous oxygen dynamics and impaired end-organ perfusion. This modular design enables the same digital twin architecture to support distinct ECMO indications without altering the underlying control formulation.

On this predictive physiological substrate, pathology-specific MPC controllers compute optimal ECMO control actions subject to explicit actuator and safety constraints. The controllers coordinate extracorporeal blood flow (Qb), sweep gas flow (Qg), and sweep gas oxygen partial pressure to regulate PaO₂, PaCO₂, and SvO₂. By solving a constrained finite-horizon optimization problem at each control step, the controller anticipates future physiological responses to metabolic disturbances and ECMO adjustments, enabling proactive rather than reactive regulation of gas exchange. The complete closed-loop architecture is illustrated in Figure 2, highlighting the bidirectional coupling between physiological prediction, oxygenator dynamics, pathology modulation, and MPC-driven control.

**Figure 2:**
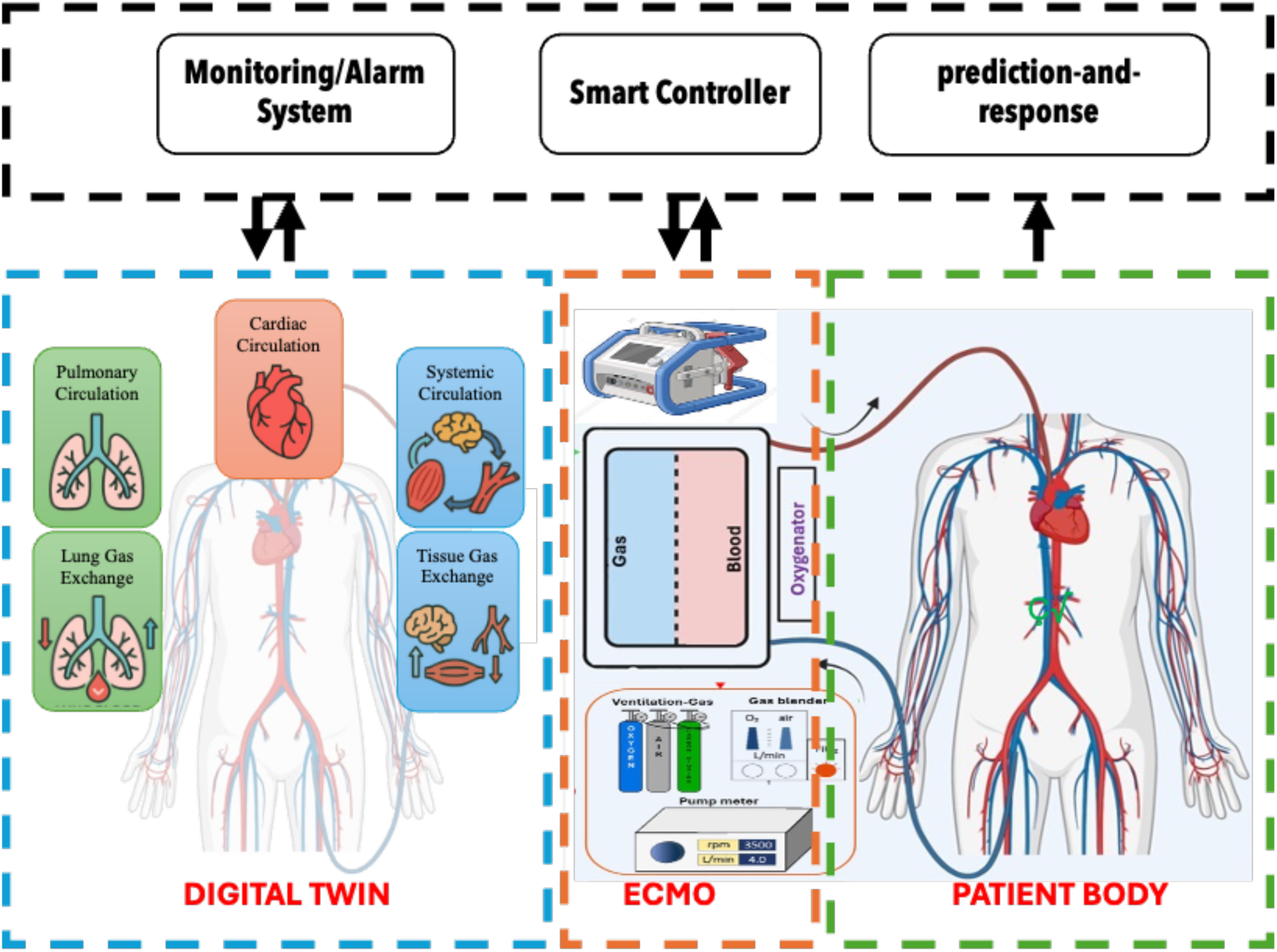
Closed-loop digital twin architecture integrating cardiopulmonary physiology, membrane oxygenator dynamics, pathology modulation, and MPC-based ECMO con- trollers.

By embedding extracorporeal life support within a predictive digital replica of patient physiology, the proposed system reframes ECMO from a manually adjusted device into a physiology-aware autonomous control framework. This architecture supports in-silico safety testing, systematic controller tuning across disease phenotypes, and future integration with bedside monitoring platforms, establishing a foundation for the clinical translation of digital twin–driven ECMO systems.

## 3. Methods

### Integrated Cardiopulmonary–ECMO Model

A closed-loop cardiopulmonary simulation framework was implemented in MATLAB/Simulink to investigate physiological responses to extracorporeal membrane oxygenation (ECMO) and to support control-oriented analysis. The model integrates cardiovascular hemodynamics, respiratory mechanics, blood–gas transport, autonomic regulation, and a membrane oxygenator into a unified dynamical system. An overview of the complete Simulink architecture is shown in Figure 3.

**Figure 3:**
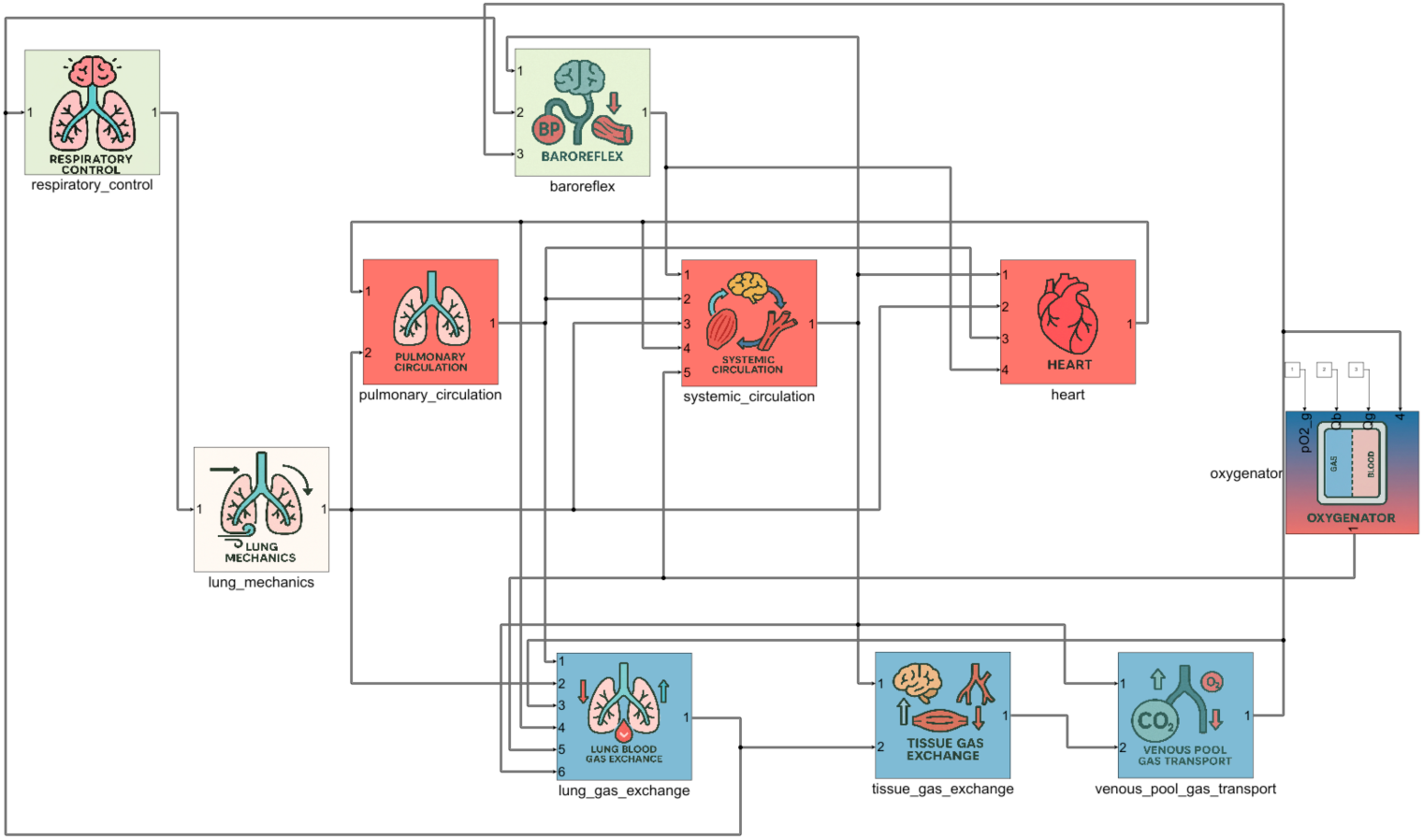
Overview of the integrated cardiopulmonary Simulink model.

The final model consists of 138 coupled differential and algebraic equations with 432 physiological and device parameters, documented in the Appendix. Systemic and pulmonary circulation were represented using a lumped-parameter (0D) hydraulic network, in which each vascular compartment is characterized by resistance (R), compliance (C), and unstressed volume (V_0_). Pressure–volume and flow relationships follow standard formulations,

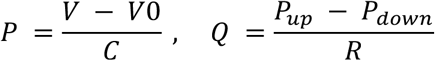

Cardiac chambers were modeled using time-varying elastance functions to generate pulsatile flow under autonomic control. Blood–gas transport was coupled to circulation via arterial, venous, and tissue gas stores, accounting for metabolic oxygen consumption and carbon dioxide production. The structure of the circulation and gas-transport subsystems is illustrated in Figures 4.

**Figure 4:**
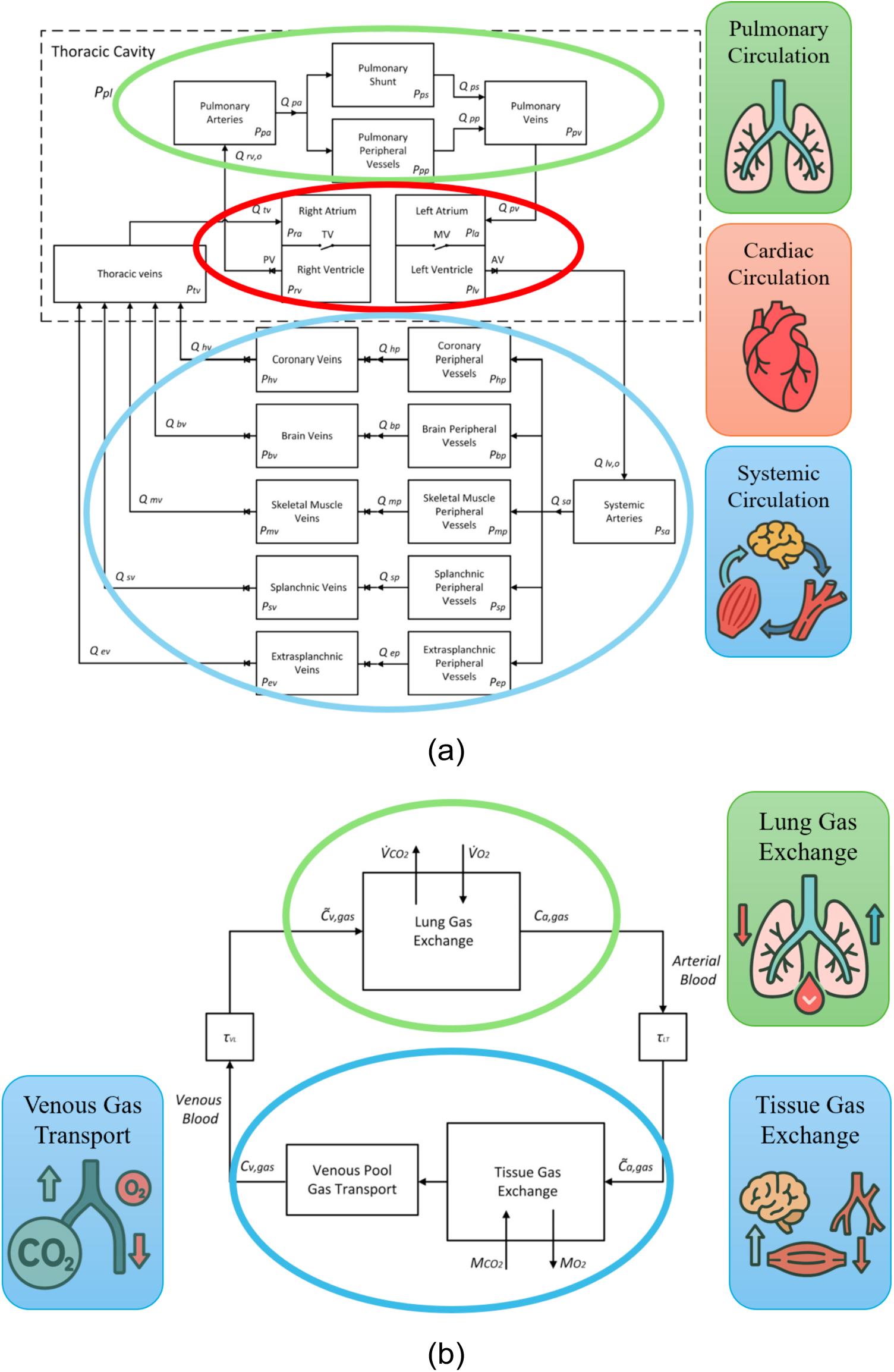
(a) Overview of the lumped-parameter flow model [51]. (b) Structure of the blood gas transport model [51].

### Oxygenator Gas-Exchange Model

The ECMO oxygenator was modeled using a multi-compartment diffusion-based formulation derived from the Hexamer–Werner framework. Blood and sweep gas phases are separated by a semi-permeable membrane, with transmembrane gas transfer for species (i) governed by Fick’s law,

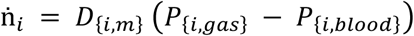

where (*D_i_*) denotes the effective membrane diffusing capacity. Blood-gas mass balance within each compartment follows

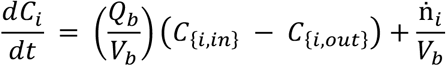

with (*Q_b_*) representing ECMO blood flow and (V) the compartment blood volume. Nonlinear oxygen and carbon dioxide dissociation relationships were used to map between partial pressure and total blood content, as shown in Figure 5 (a). The oxygenator configuration and counter-current flow arrangement are illustrated in Figure 5 (b).

**Figure 5:**
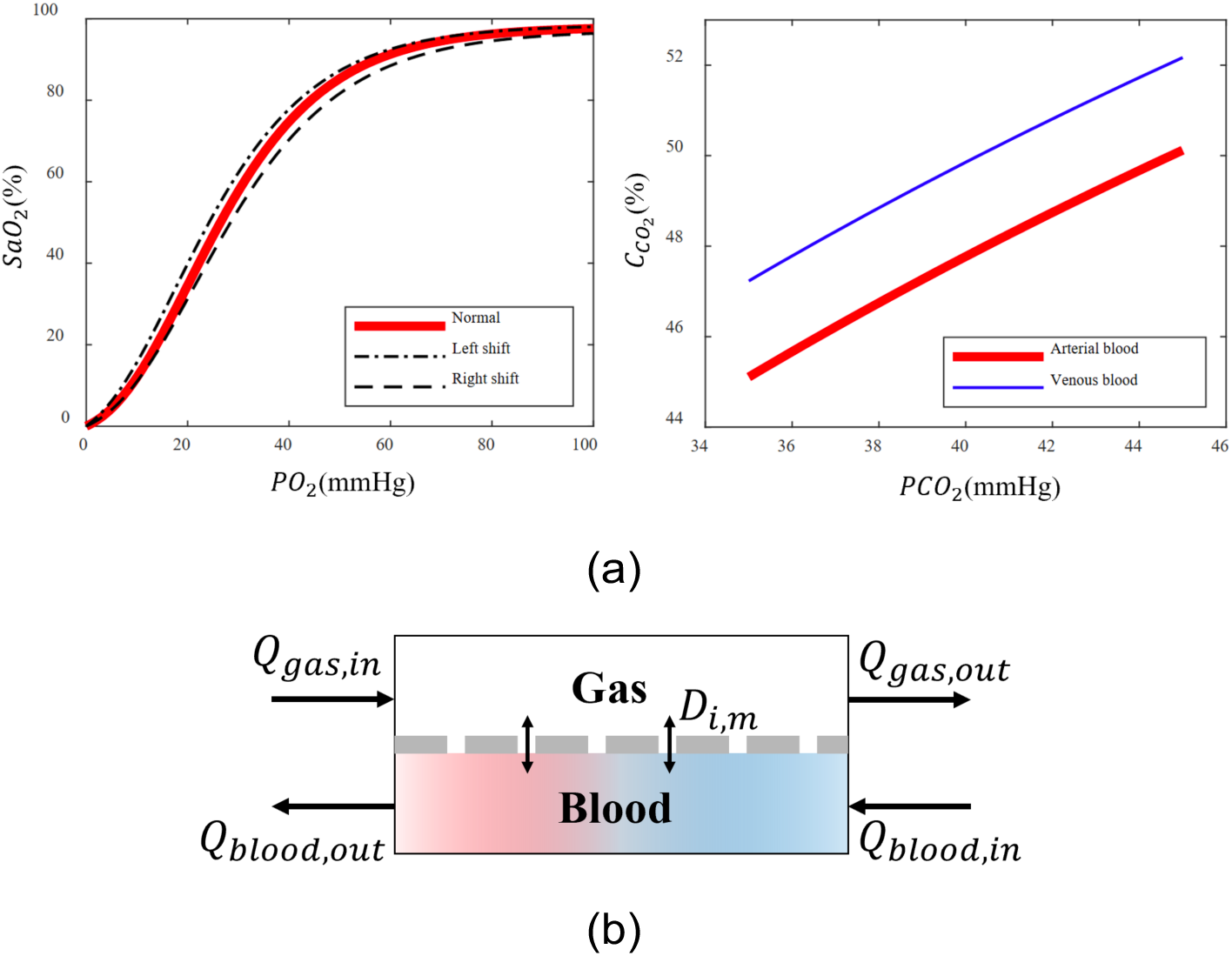
(a) Oxygen and carbon dioxide dissociation curves [2]. (b) Schematic representation of gas exchange in a membrane oxygenator [48]

### ECMO–Physiology Coupling

ECMO was introduced as a parallel circulation pathway draining blood from the thoracic venous compartment and returning oxygenated blood directly to the arterial compartment. Total arterial inflow is therefore given by

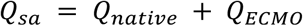

Gas concentrations from the native lung and oxygenator pathways were combined proportionally and filtered through arterial gas-store dynamics prior to systemic distribution. The coupling of flow and gas exchange within the modified circulation is illustrated in Figures 6.

**Figure 6:**
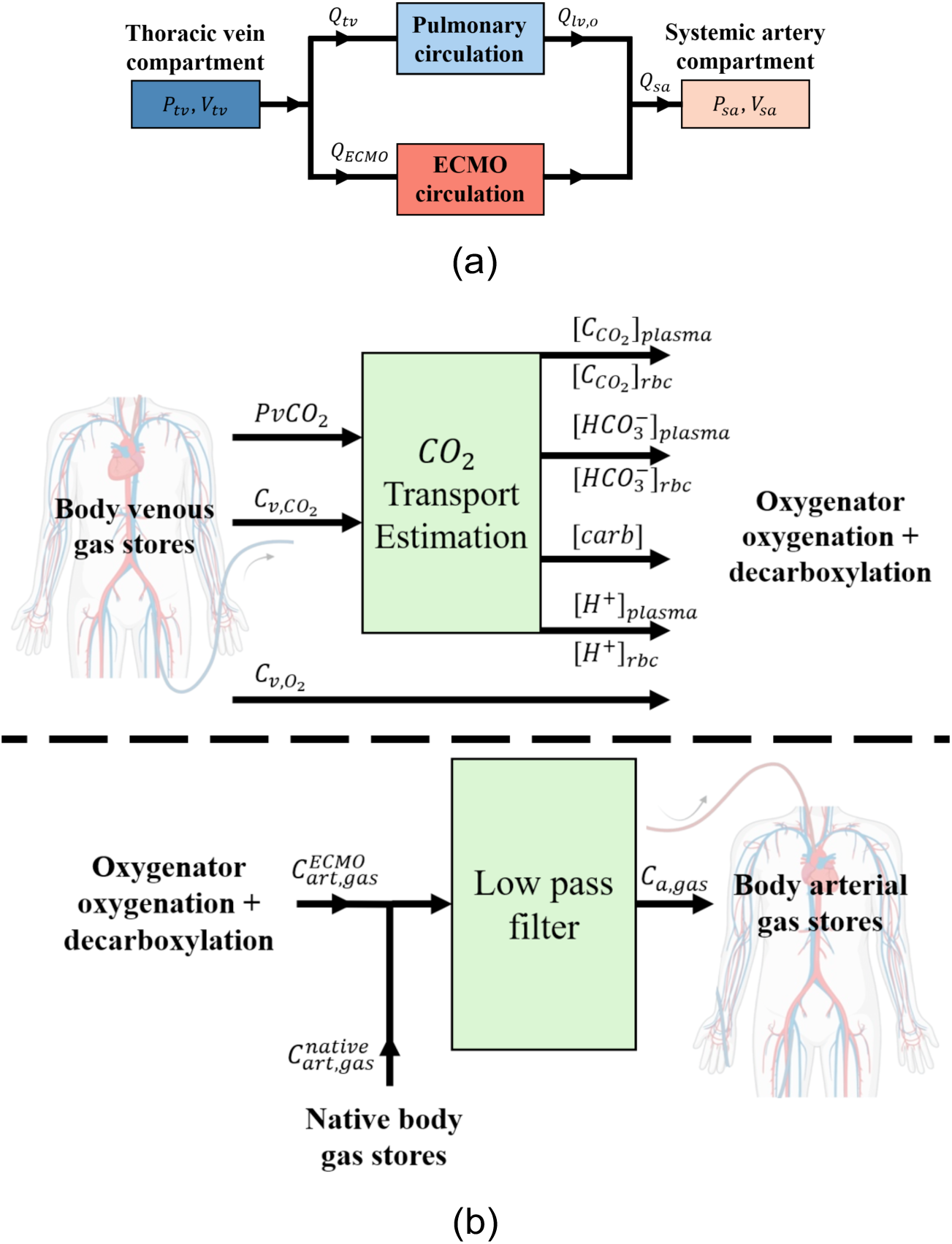
(a) Flow structure of the modified circulation model integrating native cardiac output and ECMO flow. (b) Schematic representation of the blood gas transport pathway used in the model.

### Pathophysiological Simulations

Acute respiratory distress syndrome (ARDS) was simulated by reducing lung and chest-wall compliance, increasing airway resistance and intrapulmonary shunt fraction, and impairing pulmonary gas exchange. Target arterial thresholds for *PaO*_2_ and *PaCO*_2_ followed established clinical criteria. Representative steady-state responses are shown in Figure 7 (a). Cardiogenic shock (CS) was reproduced by reducing ventricular contractility, increasing systemic vascular resistance, and elevating filling pressures, leading to reduced cardiac output and depressed mixed venous oxygen saturation (*SvO*_2_). Hemodynamic targets were selected according to ELSO guidelines, with representative results shown in Figure 7 (b).

**Figure 7:**
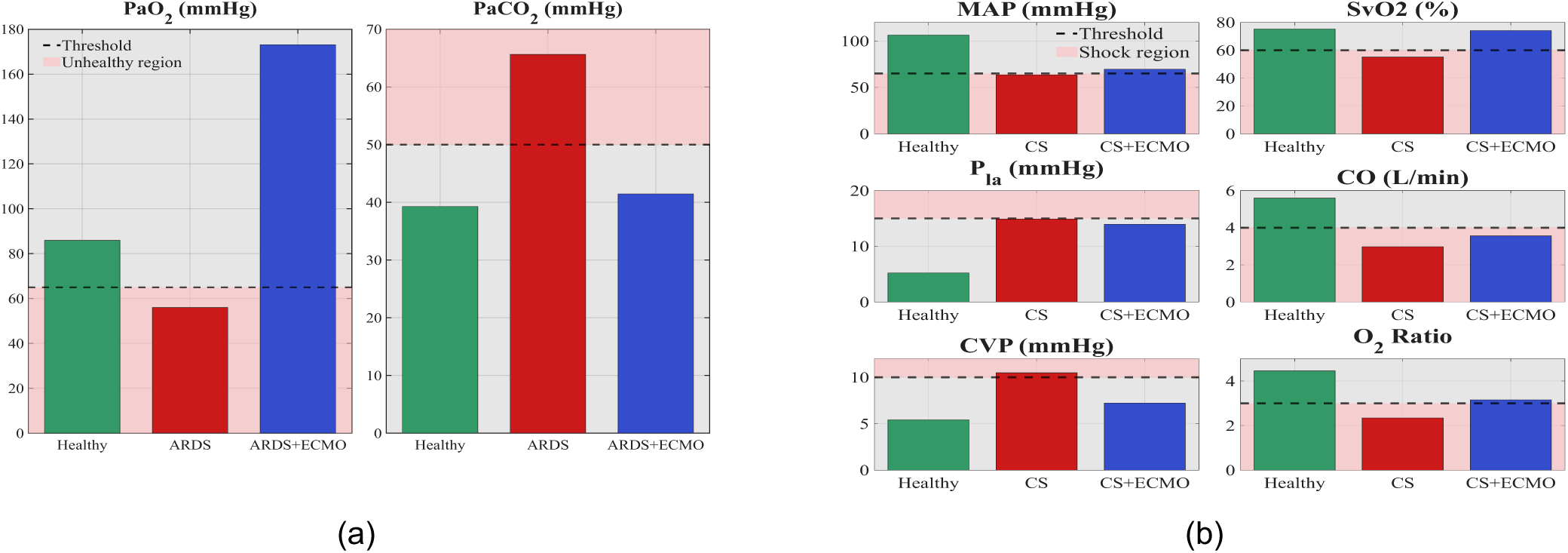
(a) Comparison of arterial oxygenation and carbon dioxide levels across Healthy, ARDS, and ARDS+ECMO conditions. (b) Hemodynamic and oxygenation comparison between Healthy, Cardiogenic Shock (CS), and CS+ECMO conditions

**Figure 8.**
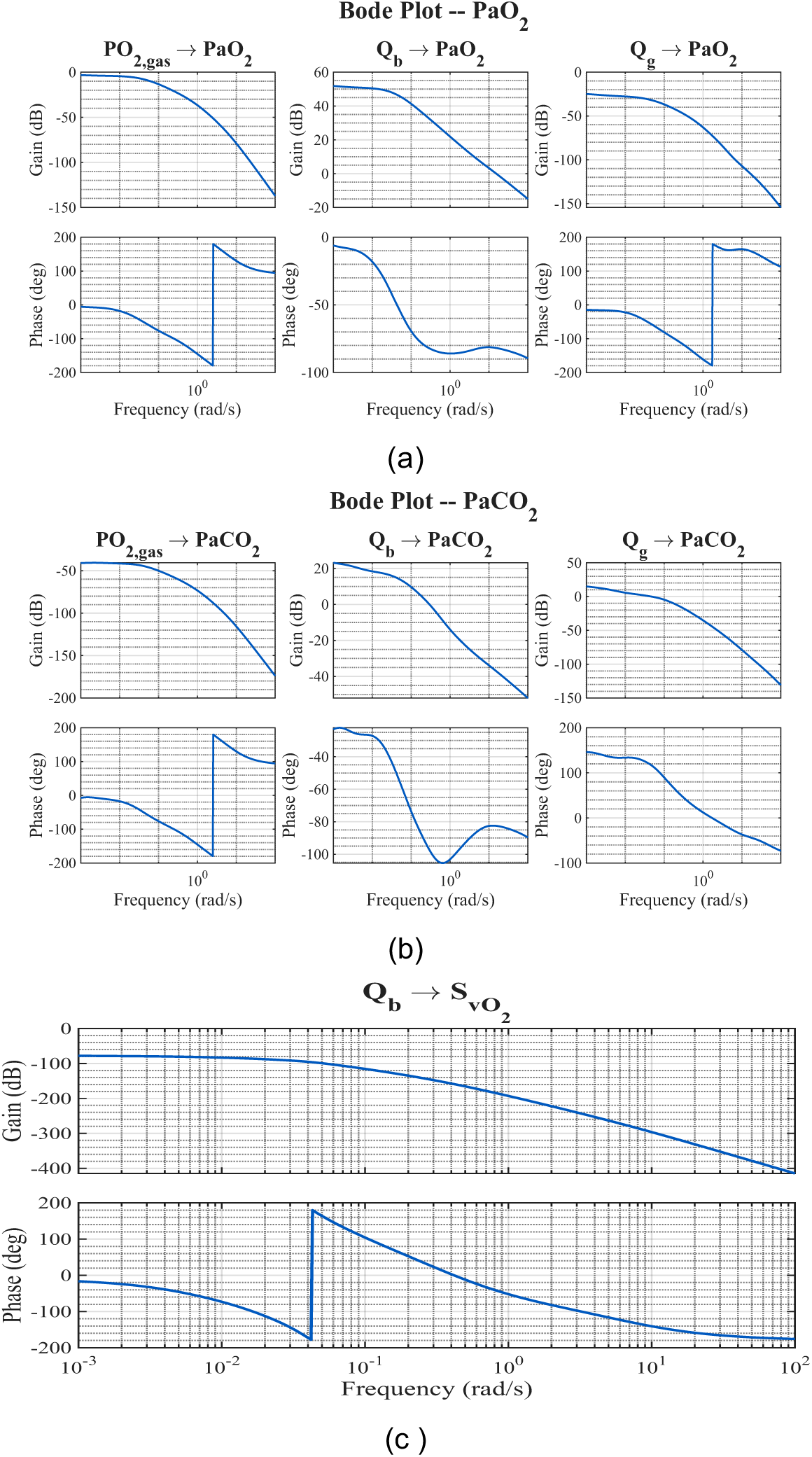
(a) Bode Plot of ARDS PaO2 Dynamics. (b) Bode Plot of PaCO2 Dynamics (c)Bode Plot of the Qb → SvO2 Dynamics.

### Linearization and Frequency-Domain Analysis

For control design, the nonlinear cardiopulmonary–ECMO model was linearized around pathology-specific steady-state operating points using a first-order Taylor expansion,

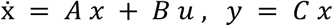

where (*x*) denotes state deviations and (*u* = [*Q_b_*, *Q_g_*, *PO*_2,*gas*)_]) represents ECMO control inputs. Frequency-domain analysis was performed to assess input–output coupling and dominant time scales governing *PaO*_2_, *PaCO*_2_, and *SvO*_2_ dynamics. Bode plots for these relationships are shown in Figures 12–14.

### Model Predictive Control Design

Pathology-specific model predictive controllers (MPCs) were designed separately for ARDS and cardiogenic shock scenarios. At each control step, a constrained quadratic optimization problem was solved,

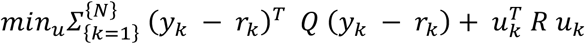

subject to the linearized system dynamics and physiological safety constraints on ECMO inputs. The manipulated variables include ECMO blood flow, sweep-gas flow, and sweep-gas oxygen partial pressure, while regulated outputs include *PaC*_2_, *PaCO*_2_, and *SvO*_2_. The MPC follows a standard receding-horizon formulation. Closed-loop tracking and disturbance-rejection performance are reported in Figures 9.

**Figure 9:**
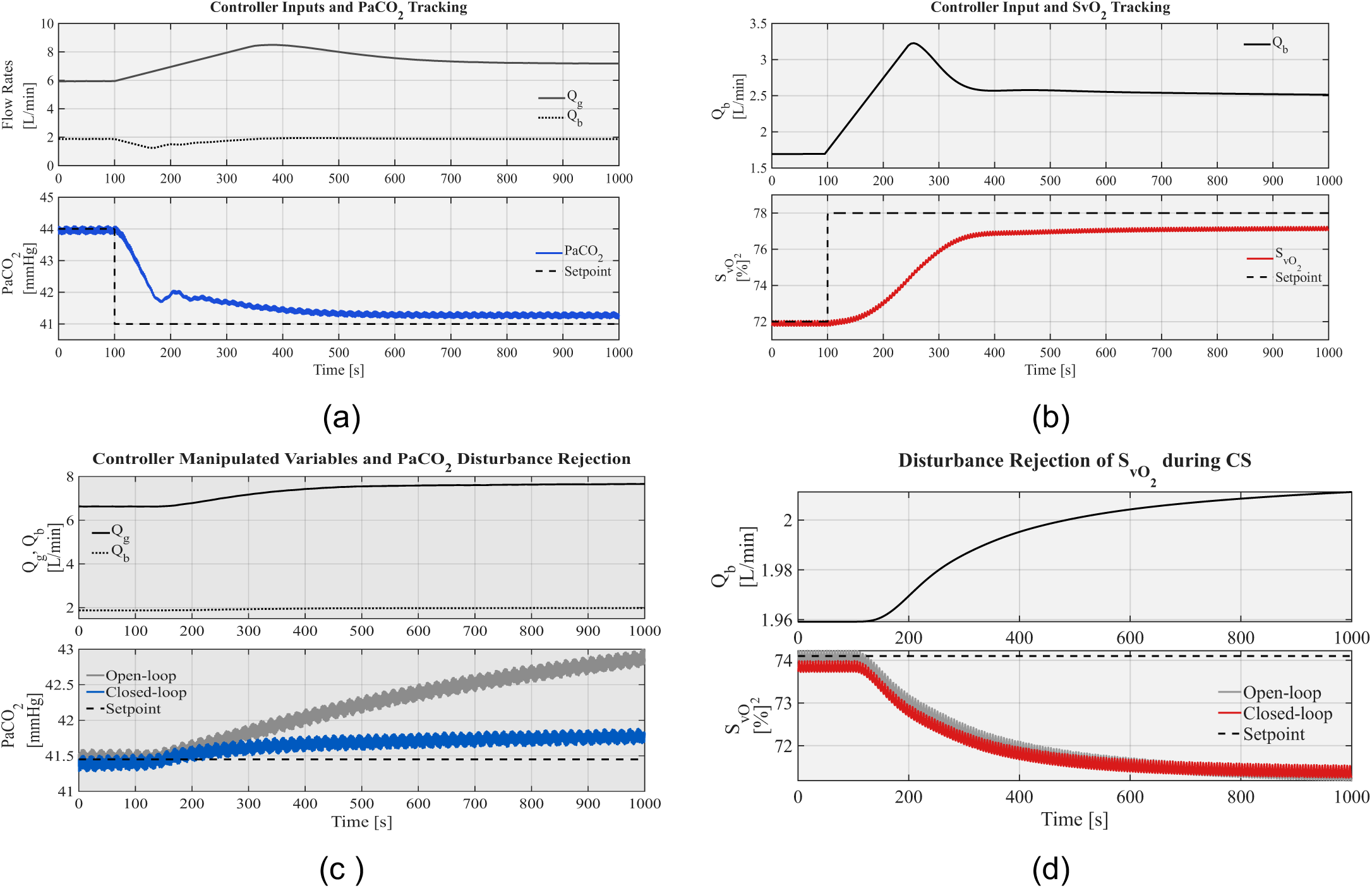
(a)Controller inputs and PaCO2 tracking during cardiogenic shock. (b) Controller input and SvO2 tracking during cardiogenic shock.(c) Controller manipulated variables and PaCO2 disturbance rejection. (d) Disturbance rejection of SvO2 during cardiogenic shock.

### Simulation Protocol and Metrics

All simulations were conducted with physiologically realistic disturbances in metabolic demand. Controller performance was evaluated using rise time, settling time, steady-state error, and disturbance attenuation metrics. Open-loop and closed-loop responses were compared to assess stabilization capability under both ARDS and cardiogenic shock conditions. All results presented in this study were generated using deterministic, reproducible computational simulations. No patient-level data, retrospective clinical records, or real-time bedside measurements were used. Model structure, parameterization, control logic, and evaluation protocols were fixed a priori to ensure transparency and reproducibility of all reported findings.

## 4. Results

### Reproduction of ARDS and Cardiogenic Shock Physiology

Prior to controller activation, the integrated cardiopulmonary, ECMO model was evaluated to verify that pathology-specific parameterization reproduced hallmark physiological features of acute respiratory distress syndrome (ARDS) and cardiogenic shock (CS). Under ARDS conditions, reduced lung compliance and elevated intrapulmonary shunt fraction led to attenuated alveolar volume excursions and impaired pulmonary gas exchange. Figure 10(a) shows a marked reduction in alveolar volume amplitude compared with healthy baseline conditions. These mechanical alterations resulted in sustained arterial hypoxemia and hypercapnia, with *PaO*_2_ and *PaCO*_2_ deviating beyond clinically acceptable thresholds. As summarized in Fig. 10(b), ECMO support partially restored arterial oxygenation and carbon dioxide levels toward physiological ranges, confirming correct coupling between extracorporeal and native gas-exchange pathways. In cardiogenic shock simulations, depressed ventricular contractility and increased systemic vascular resistance produced reduced cardiac output, elevated central venous pressure, and decreased mixed venous oxygen saturation (*SvO*_2_). Steady-state hemodynamic and oxygenation variables shown in Fig. 11 align with established cardiogenic shock criteria, confirming successful reproduction of a hemodynamic-dominant failure state prior to controller engagement.

**Figure 10:**
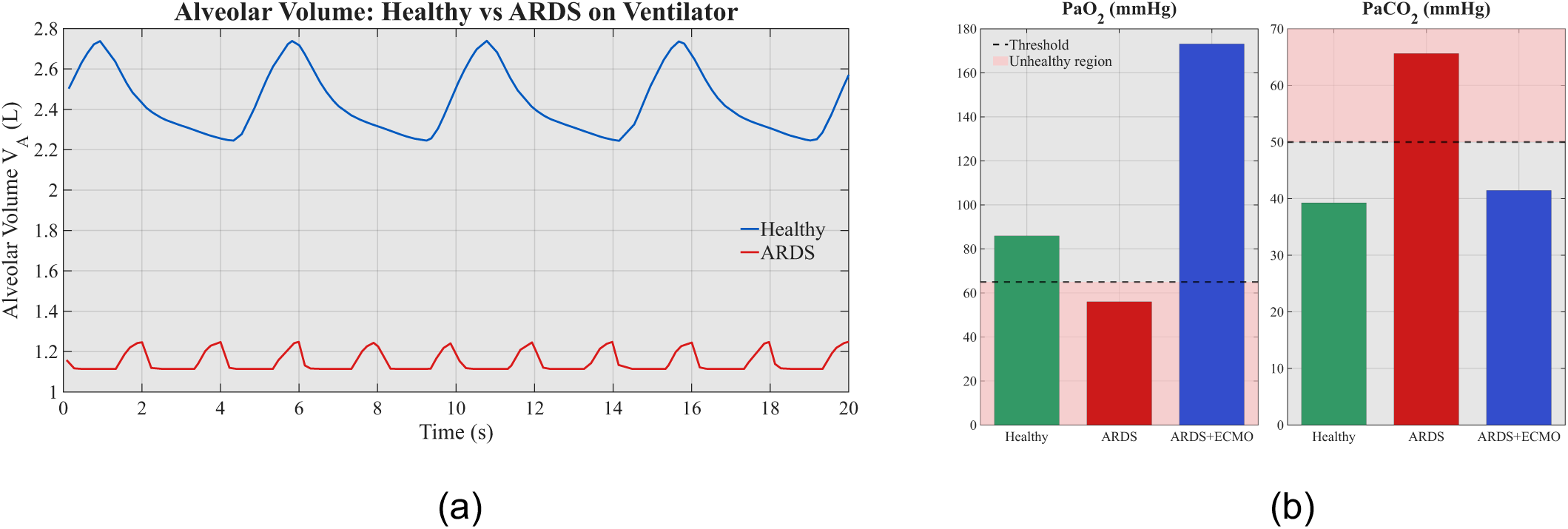
(a) Comparison of healthy and ARDS alveolar volume (VA) over a 20-second interval. The healthy waveform is shown in blue and the ARDS waveform in red. (b) Comparison of arterial oxygenation and carbon dioxide levels across Healthy, ARDS, and ARDS+ECMO conditions.

**Figure 11:**
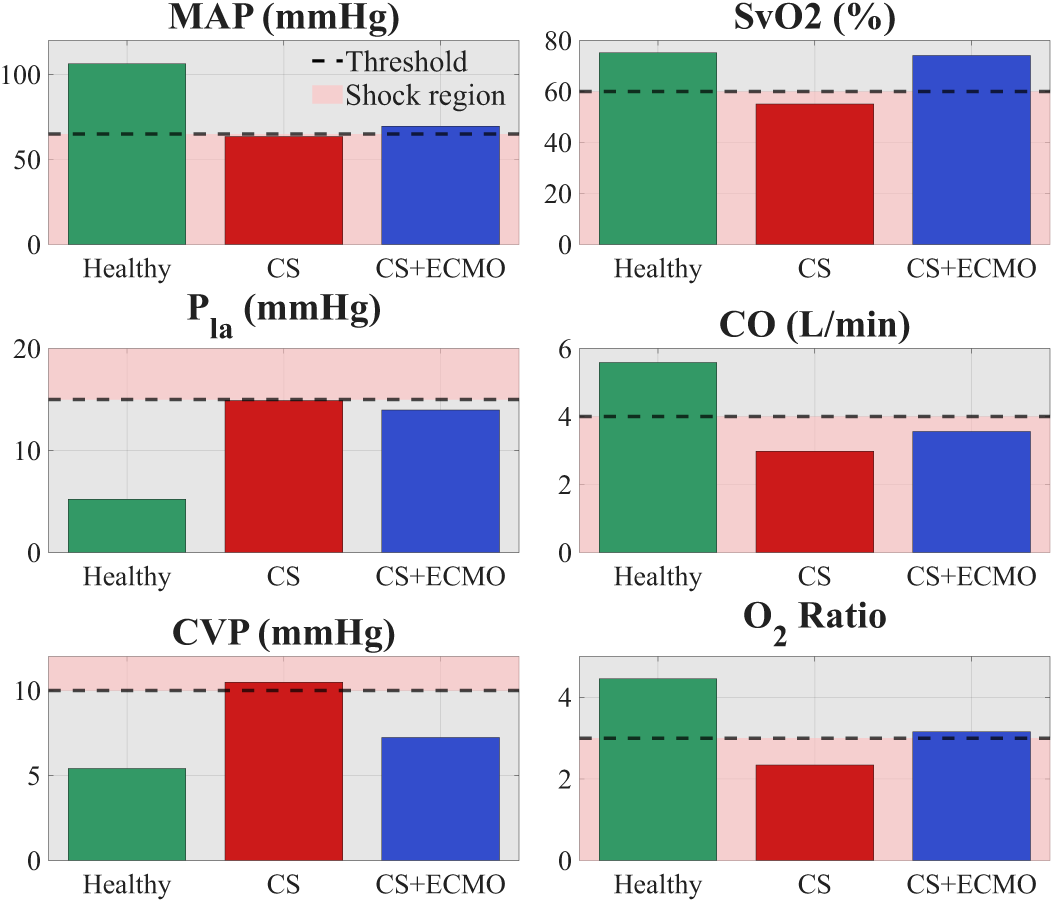
Hemodynamic and oxygenation comparison between Healthy, Cardiogenic Shock (CS), and CS+ECMO conditions.

In cardiogenic shock simulations, depressed ventricular contractility and increased systemic vascular resistance produced reduced cardiac output, elevated central venous pressure, and decreased mixed venous oxygen saturation (*SvO*_2_). Steady-state hemodynamic and oxygenation variables shown in Fig. 11 align with established cardiogenic shock criteria, confirming successful reproduction of a hemodynamic-dominant failure state prior to controller engagement.

### Open-Loop ECMO Sensitivity and Dynamic Response

Open-loop simulations were conducted to characterize intrinsic ECMO–physiology interactions and dominant system time scales. In ARDS conditions, step increases in ECMO blood flow resulted in rapid elevation of PaO₂ with relatively minor effects on PaCO₂ (Fig. 12a). Conversely, sweep-gas flow primarily modulated PaCO₂, enabling efficient carbon dioxide removal with limited impact on oxygenation (Fig. 12b). Adjustments to sweep-gas oxygen partial pressure produced proportional changes in PaO₂ while exerting negligible influence on PaCO₂ (Fig. 12c). These responses demonstrate partial decoupling between oxygenation and ventilation control channels, while revealing residual cross-coupling inherent to cardiopulmonary physiology. In cardiogenic shock simulations, step increases in ECMO blood flow improved SvO₂ and mean arterial pressure over significantly slower time scales (Fig. 12d), reflecting the dominant influence of tissue oxygen stores and circulatory transport delays.

**Figure 12:**
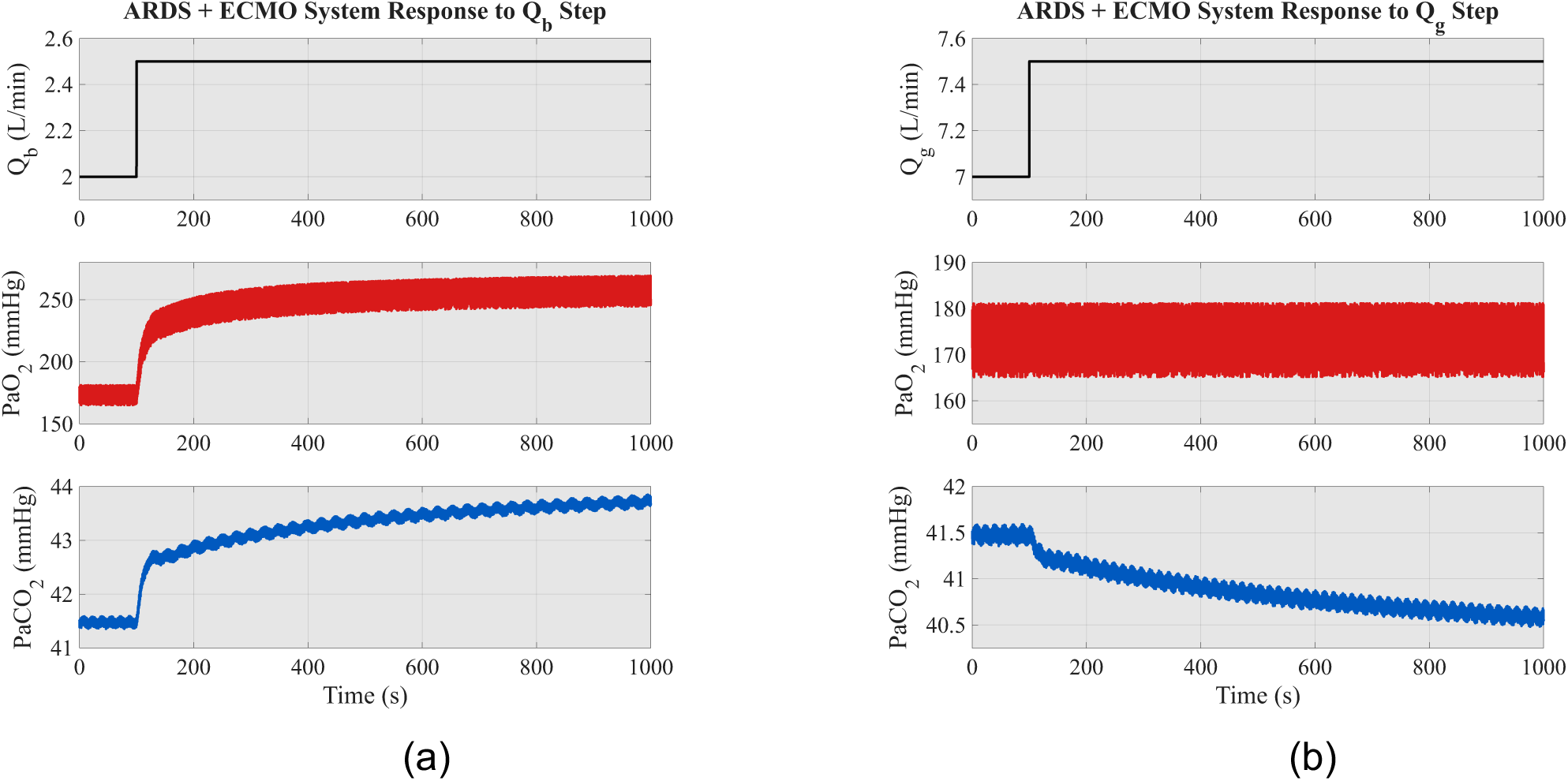
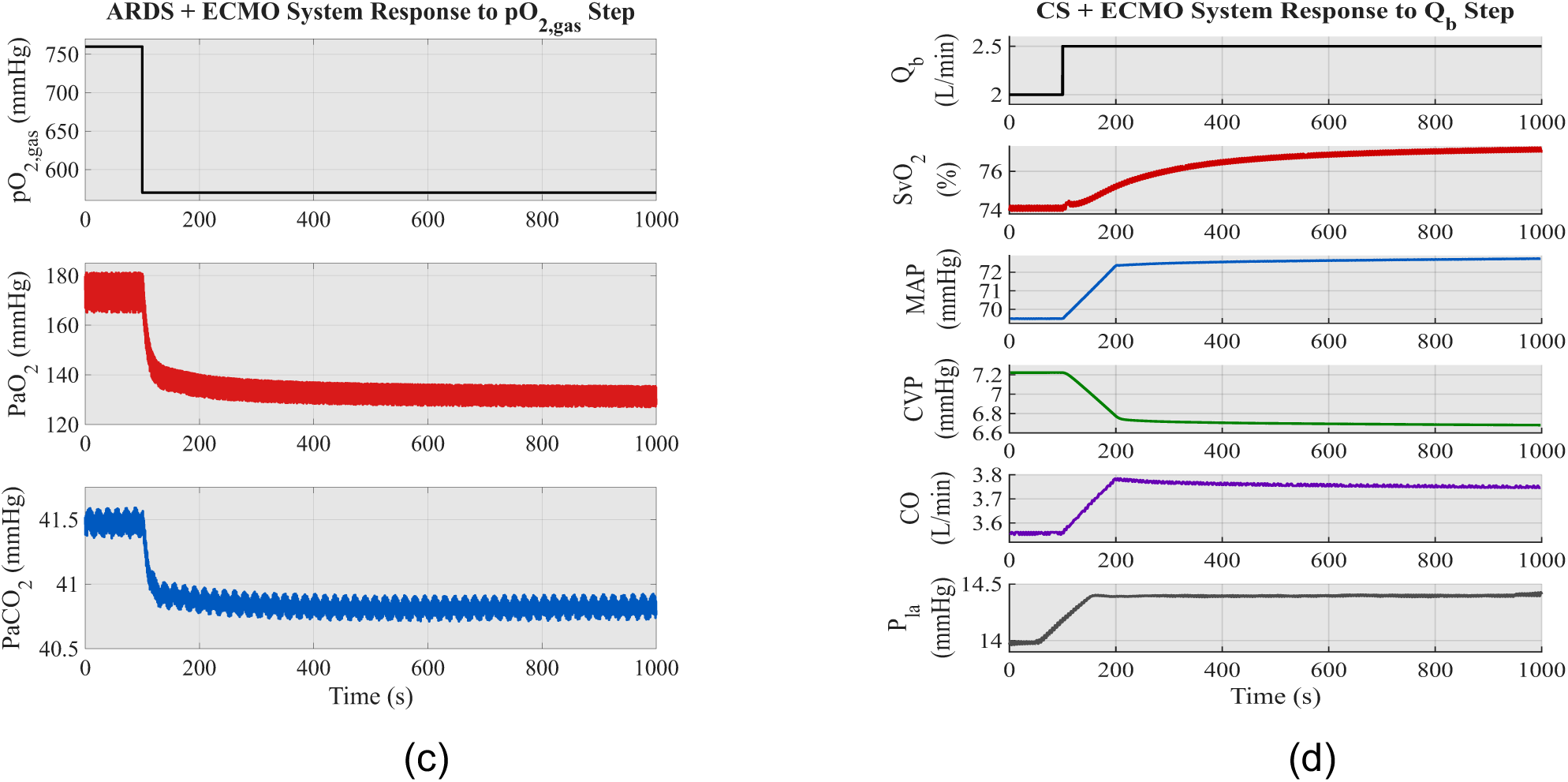
(a) ARDS + ECMO response to a step increase in ECMO blood flow (*Q*_b_). (b) ARDS + ECMO response to a step increase in oxygenator sweep-gas flow rate (*Q*_g_). (c) Controller manipulated variables and PaCO2 disturbance rejection. (d) ARDS + ECMO response to a step change in oxygenator sweep gas oxygen partial pressure (*pO*_2,*gas*_).

### Closed-Loop Control Performance in ARDS

With model predictive control enabled, the ARDS-specific controller achieved rapid and stable regulation of arterial gas tensions. Following reference changes, PaCO₂ converged to target values with minimal overshoot and short settling times (Fig. 9a). PaO₂ regulation exhibited similarly stable behavior without violation of actuator constraints. Disturbance rejection performance was assessed by increasing metabolic CO₂ production. Closed-loop control substantially attenuated PaCO₂ excursions compared with open-loop operation (Fig. 9c), maintaining arterial levels within physiologically acceptable limits. Quantitative performance metrics, including rise time, settling time, and steady-state error, confirmed robust tracking and effective disturbance rejection across tested scenarios.

### Closed-Loop Control Performance in Cardiogenic Shock

In cardiogenic shock simulations, the MPC successfully regulated mixed venous oxygen saturation through coordinated modulation of ECMO blood flow. SvO₂ tracking following step reference changes demonstrated stable convergence without inducing hemodynamic instability (Fig. 9b). When metabolic oxygen consumption was increased, closed-loop control reduced but did not fully eliminate SvO₂ deviations (Fig. 9d). This limitation reflects intrinsic physiological time constants associated with venous oxygen transport and tissue oxygen buffering rather than controller instability. Despite these constraints, closed-loop operation consistently outperformed open-loop behavior, improving systemic oxygen delivery while respecting hemodynamic and actuator safety limits.

Across both ARDS and cardiogenic shock scenarios, the closed-loop digital twin framework achieved stable regulation of clinically relevant gas-exchange and perfusion variables under realistic disturbances. Control performance was pathology dependent: ARDS regulation benefited from relatively fast gas-exchange dynamics, whereas cardiogenic shock control was constrained by slow circulatory transport and venous oxygen kinetics. Nevertheless, in all cases, model predictive control improved physiological stability compared with open-loop ECMO operation.

## 5. Discussion

The proposed system is intended as clinical decision-support software, with fully autonomous control evaluated exclusively in silico prior to any prospective clinical or preclinical deployment. This study also demonstrates the technical and physiological feasibility of fully closed-loop ECMO automation using a digital twin–driven model predictive control framework. Unlike prior approaches that rely on simplified gas-exchange surrogates or single-loop proportional–integral strategies, the proposed system embeds extracorporeal life support within a coupled cardiopulmonary digital twin, enabling predictive regulation based on physiological state evolution rather than purely reactive feedback. In ARDS simulations, gas-exchange dynamics dominated by diffusion limitation and intrapulmonary shunt were robustly stabilized using multivariable predictive control. Across all tested perturbations, PaO₂ and PaCO₂ were rapidly restored to their target ranges with minimal overshoot and negligible steady-state error. These results indicate that oxygenation and ventilation can be effectively decoupled from abrupt extracorporeal and metabolic disturbances, a capability that is not achievable with conventional manual titration strategies. Importantly, all actuator trajectories remained within predefined safety limits, demonstrating that aggressive stabilization does not require unsafe operating conditions.

A practical clinical translation pathway for this framework is a staged deployment that begins with human-in-the-loop decision support rather than unsupervised autonomy. In an initial implementation, the digital twin would run in parallel with standard ECMO management, ingesting routinely available bedside measurements (e.g., blood gas values when available, continuous SpO₂, end-tidal CO₂ when applicable, pump settings, and hemodynamic signals) to generate short-horizon forecasts and recommended parameter adjustments for *Q_b_*, sweep gas flow, and sweep gas oxygen fraction. Recommendations could be presented to clinicians with explicit predicted impacts on *PaO*_2_, *PaCO*_2_, and *SvO*_2_ and with constraint-aware “do-not-exceed” boundaries to preserve safety. This approach aligns with contemporary ICU workflows, where adjustments are often made under time pressure and uncertainty, and where delays between physiologic deviation and corrective action can be clinically consequential.

For autonomy-assisted operation, safety must be treated as a first-class objective. The present framework already incorporates actuator limits and constrained optimization; however, translation will require additional layers including measurement-noise handling, sensor fault detection, actuation latency modeling, and explicit fail-safe behavior (e.g., automatic reversion to a conservative baseline setting or alert-only mode). Importantly, the system is best conceptualized as **clinical decision support software** that can be evaluated under prospective protocols before any escalation toward higher levels of automation. Future work should therefore focus on (i) validating model fidelity against bench oxygenator testing and animal or clinical retrospective datasets, (ii) implementing online parameter estimation to adapt to evolving patient physiology and oxygenator aging, and (iii) assessing workflow integration, clinician trust, and safety outcomes under simulated and controlled deployment scenarios.

In contrast, cardiogenic shock control revealed a fundamentally different physiological control regime dominated by circulatory transport delays and impaired systemic oxygen delivery. The slower convergence of SvO₂ observed under cardiogenic shock reflects intrinsic venous oxygen transport time constants and tissue oxygen buffering rather than limitations of the control algorithm itself. Nevertheless, the closed-loop system consistently preserved systemic oxygen delivery without inducing hemodynamic instability or secondary gas-exchange dysregulation. This pathology-dependent divergence in control behavior directly supports the central premise of this work: ECMO automation cannot rely on a single universal control law but must instead adapt control objectives and tuning to the underlying physiological failure mode.

From a clinical automation perspective, the proposed framework transforms ECMO from a predominantly reactive support modality into a continuously optimized physiological control system. The ability to autonomously compensate for fluctuations in metabolic demand, extracorporeal flow disturbances, and nonlinear gas-transfer dynamics in real time addresses a critical safety and workload bottleneck in contemporary ECMO practice. This capability is particularly relevant in high-acuity environments, where delayed blood-gas sampling and operator burden may expose patients to prolonged episodes of hypoxemia or hypercapnia. Beyond immediate control performance, the digital twin architecture provides a foundation for patient-specific adaptation. By embedding physiology, pathology, and extracorporeal mechanics within a unified predictive framework, the system enables individualized controller tuning, in-silico therapy testing, and future integration with adaptive or learning-based control strategies. These capabilities are essential for advancing ECMO automation beyond static rule-based systems toward intelligent, data-driven life-support platforms.

This study has several important limitations. First, all results are derived from in-silico validation using a lumped-parameter cardiopulmonary model. While this approach captures the dominant hemodynamic and gas-exchange dynamics relevant to ECMO control, it does not resolve spatial heterogeneity in pulmonary ventilation–perfusion mismatch or microvascular perfusion. Second, the framework does not account for cannula recirculation, thrombus formation, oxygenator aging, or sensor drift, all of which may influence real-world ECMO performance. Measurement noise, actuation latency, and blood-gas analyzer delays were also not explicitly modeled. Third, controller tuning was performed around representative pathology-specific operating points rather than adapting continuously to evolving disease trajectories. Although sufficient to demonstrate feasibility and robustness, clinical translation will require online adaptation and parameter estimation. Finally, no animal or human validation has yet been conducted. Translation to clinical practice will therefore require staged preclinical evaluation followed by carefully regulated prospective studies before autonomous ECMO deployment can be considered.

## 5. Conclusion

We present a closed-loop digital twin framework for autonomous ECMO control under two fundamentally distinct forms of cardiopulmonary failure: acute respiratory distress syndrome and cardiogenic shock. By integrating detailed physiological modeling with pathology-specific model predictive control, the proposed system achieves stable, robust, and safe regulation of oxygenation, ventilation, and systemic oxygen delivery under a wide range of disturbances. The results demonstrate that ECMO automation is technically feasible within strict physiological and actuator safety constraints while accommodating fundamentally different disease dynamics. This work establishes a methodological foundation for the development of intelligent extracorporeal life-support systems and supports the progression from operator-driven ECMO toward predictive, patient-specific, and autonomy-assisted critical care.

## Ethics Statement

This study was conducted exclusively using computational simulations. No human participants, animal subjects, or identifiable patient data were used. Institutional review board approval and informed consent were there- fore not required.

## Data Availability

All simulation models, controller implementations, and analysis scripts used in this study will be made publicly available for academic research purposes through the Nezami Lab GitHub repository: https://github.com/Nezami-lab/ECMO_Control. Patient-level clinical data are not applicable, as this study was conducted entirely in silico.

